# Dementia Care Specialists Perspectives of Diagnosis and Early Psychosocial Care: A Qualitative Analysis of Focus Groups in Two Large Academic Medical Centers

**DOI:** 10.1101/2024.12.04.24316485

**Authors:** Amelia J. Hicks, Julie Brewer, Nina Ahmad, Talea Cornelius, Robert A. Parker, Kristen Dams-O’Connor, Bradford Dickerson, Christine Ritchie, Ana-Maria Vranceanu, Sarah M. Bannon

**Affiliations:** Brain Injury Research Center, Department of Rehabilitation and Human Performance, Icahn School of Medicine at Mount Sinai Hospital, New York, NY; Center for Health Outcomes and Interdisciplinary Research, Department of Psychiatry, Massachusetts General Hospital and Harvard Medical School, Boston, MA; Center for Behavioral Cardiovascular Health, Columbia University Irving Medical Center, New York, NY; Biostatistics Center, Massachusetts General Hospital and Harvard Medical School, Boston, MA; Frontotemporal Disorders Unit, Departments of Neurology and Psychiatry, Massachusetts General Hospital and Harvard Medical School, Boston, MA; Mongan Institute Center for Aging and Serious Illness and the Division of Palliative Care and Geriatric Medicine, Department of Medicine, Massachusetts General Hospital and Harvard Medical School, Boston, MA

**Keywords:** dementia care specialist specialists, dementia diagnosis, post-diagnosis support, focus group

## Abstract

**Background and Objective:** Alzheimer’s disease and related dementias (ADRDs) are progressive conditions that substantially impact individuals and families. Timely diagnosis and early support are critical for long-term adjustment. However, current dementia care models do not meet needs of patients and families. Dementia care specialists treating individuals with dementia offer unique insight into care needs of diverse groups of patients, families, and healthcare systems that can be used to identify opportunities to improve care.

To understand dementia care specialists’ impressions of factors impacting ADRD diagnosis and post-diagnosis support. We aimed to identify factors that impact: (1) timely and accurate diagnosis, (2) diagnostic disclosure and provision of post-diagnosis support, and (3) patient and care-partner adjustment after diagnosis.

**Research Design and Methods:** We recruited dementia care specialists treating persons living with dementia (n=19) from two academic medical centers. Participants completed 60-minute qualitative focus groups or individual interviews. Data were analyzed using a hybrid inductive-deductive approach to thematic analysis.

**Results:** We identified subthemes within three overarching a-priori determined themes. Participants highlighted the presence of delays in referrals, time constraints, specialist discomfort, and lack of training as factors impacting the timeliness and accuracy of diagnosis. They also highlighted information needed in disclosure visits, ways of coordinating care, and identifying early support needs. Finally, participants highlighted factors impacting adjustment including families’ insight and acceptance, distress, and available resources.

**Discussion and Implications:** Our study highlights the challenges dementia care specialist specialists face in delivering early support for individuals and families impacted by ADRDs and suggests avenues for revising existing care models.

## INTRODUCTION

Alzheimer’s disease and related dementias (ADRDs) are progressive and disabling conditions that have substantial impact on the health, quality of life, and relationships of individuals and their families (Edwards et al., 2018; Gellert et al., 2018; Laakkonen et al., 2008; Petty et al., 2018). After diagnosis, emotional distress is common and interdependent (i.e., correlated outcomes and mutual influence)(Balhara et al., 2012; Shaffer et al., 2017), for both patients (Aminzadeh et al., 2007; Laakkonen et al., 2008; Petty et al., 2018) and their family care partners (Freedman et al., 2022; Harris et al., 2021; Laakkonen et al., 2008; Ma et al., 2018; Sheehan et al., 2021)—together considered a unit or *dyad*. Progressive symptoms of ADRDs (e.g., forgetfulness, communication challenges, changes in behavior and personality) can heighten this emotional distress in patient and care partner dyads, with spouses experiencing the greatest emotional impact (Edwards et al., 2018; Gellert et al., 2018; Laakkonen et al., 2008; Petty et al., 2018). Addressing emotional distress in dyads early is important given its prevalence, interdependence, the difficulty of addressing emotional distress when it becomes chronic, and the documented impact of distress on ADRD symptoms and long-term care (Campbell, 2009; Stall et al., 2019).

The 2022 World Alzheimer Report was dedicated to highlighting needs for advanced post-diagnosis support for patients and care partners (Gauthier et al., 2022) As the report mentions, patients’ and care-partners’ emotional and practical adjustment to ADRDs are influenced by: 1) timely and accurate diagnosis of ADRDs, 2) person-centered diagnostic disclosure, and 3) early resource provision (Gauthier et al., 2022). Unfortunately, diagnostic delays remain common and contribute to heightened emotional distress (Lin et al., 2021; L. Robinson et al., 2005; van Vliet et al., 2013; Volpe et al., 2020) and can impact the availability and quality of post-diagnosis support and long-term care. Delays in diagnosis are related to patient and family beliefs about the cause of symptoms and clinicians lack of confidence and discomfort communicating a diagnosis of ADRDs, particularly in generalist settings (e.g., primary care) (Gauthier et al., 2022).

The manner in which a diagnosis is communicated can also impact adjustment to ADRDs (Gauthier et al., 2022). Standards for delivering a dementia diagnosis exist, but are not widely implemented and did not consider patient and care partner input until this past year (Armstrong et al., 2024). Recent consensus guidelines highlight the importance of delivery using compassion and empathy, asking questions about diagnosis preference, providing practical strategies and resources, and a written plan to manage symptoms and prepare for the future (Armstrong et al., 2024). While these guidelines can provide clinicians with a road map to navigate diagnosis disclosure, it is unclear what factors are currently barriers and facilitators to more person-centered diagnosis disclosure.

Psychoeducational support can also provide patients and their care-partners with opportunities to learn about the diagnosis and symptoms, discuss role adjustments, increase support networks, and begin important legal, financial, accommodation and caregiving planning while individuals have the cognitive capacity to advocate for their wishes (Bailey et al., 2019; Rasmussen & Langerman, 2019). However, these potential benefits are curtailed by the widespread service gaps that exist immediately after diagnosis (Low et al., 2023). Approximately 64% of individuals surveyed across studies reviewed in the 2022 Alzheimer World Report indicated that after diagnosis disclosure they did not receive information on their stage of dementia and expected progression of symptoms, available pharmacological and non-pharmacological interventions, and ways of planning for the future (Gauthier et al., 2022). People living with dementia and care-partners express broad dissatisfaction with care and unmet support needs around the time of diagnosis (Walrath & Lawlor, 2019).

In general, national and international care models and policy guidelines have had minimal effect on improving peri-and post-diagnosis dementia care for patients and care partners (Low et al., 2023; Parker et al., 2020). Alternative models of care have been proposed that suggest a “tiered” support system that includes both generalist and specialist care (Low et al., 2023). Some progress has been made to provide tiered support specialist care settings such as the UCLA Alzheimer’s and Dementia Care (UCLA ADC) program. This program offers system-wide comprehensive dementia care management using nurse practitioners that act as dementia care managers, and has led to clinical benefits for patients and care partners, with some early evidence of successful dissemination to other hospital systems (Reuben et al., 2019, 2022). While this program has improved person-centered care and some early resource provision, gaps remain in early psychosocial support that patients and care partners can participate in simultaneously, and for support that adopts a dyadic or family-focused lens.

Healthcare specialists providing dementia care are ideally suited to offer important perspectives on care gaps and opportunities. Indeed, they are exposed to real-world day-to-day challenges impacting the delivery of optimal care to diverse groups of dyads navigating ADRDs within complex healthcare systems. Therefore, the present study has the primary objective of understanding dementia care specialist specialists’ perceptions of factors impacting dementia diagnosis and post-diagnosis support for patient-care partner dyads. We aimed to examine three broad areas, including (1): factors that impact timely and accurate diagnosis, (2) factors that impact diagnostic disclosure and the provision of post-diagnosis support, and (3) factors impacting patient and care-partner adjustment after diagnosis. A more detailed understanding of dementia care specialists’ perceptions of factors impacting the quality of care and early adjustment can directly inform the refinement and dissemination of early care models to feasibly meet the needs of the growing population of dyads impacted by ADRDs.

## MATERIALS AND METHODS

### Participants and Procedures

All study procedures were approved by the Massachusetts General Hospital (2022P001510) and the Icahn School of Medicine at Mount Sinai (22-01623) and Institutional Review Boards. We adhered to COREQ reporting guidelines for qualitative studies (Booth et al., 2014) and prospectively described the protocol for the study prior to data collection (Bannon et al., 2023). We recruited specialist dementia care specialists through convenience sampling from two large academic medical centers: 1) Massachusetts General Hospital and 2) Mount Sinai Hospital. Recruitment occurred in relevant departments (e.g., Neurology, Geriatric Medicine, Psychiatry) and focused on clinics and programs dedicated to the treatment of ADRDs. Participants were recruited via flyers, presentations, word of mouth referrals, and department listservs. Individuals were eligible for inclusion if they were: (1) adults (18+ years old) fluent in English, (2) willing and able to participate in a video focus group or interview, and (3) employed in a position involving direct clinical care or caregiver support for patients with dementia caused by ADRDs. A total of 21 interested individuals contacted the study team via email or by completing a screening survey on the REDCap survey platform. Two individuals were deemed ineligible on screening (research staff with no clinical role). We continued recruitment until we achieved thematic saturation on a-priori determined study domains (Boddy, 2016), which resulted in a final sample of *N*=19 participants. We obtained informed consent electronically from all participants prior to the start of qualitative focus groups and interviews. Interviews were completed between November 2022 and June 2023. No repeat interviews were completed.

Data collection was conducted separately for each of the two hospital sites to allow for potential differences by setting. Each participant completed a single 60-minute focus group or individual interview (based on scheduling availability) with a member of the study team. Interviews were semi-structured in nature and followed an interview guide (see Supplemental Materials) that was collaboratively developed by the multidisciplinary study team (clinical psychologists, geriatricians, neurologists, neuropsychologists, neuroscientists). The interview guide included open ended questions organized into domains focused on: (a) factors impacting timely and accurate diagnosis, (b) perceptions of care and support for dyads early after ADRD diagnoses, and (c) perceptions of factors impacting individual and care-partner adjustment (see Supplemental Materials). We asked participants to respond to general questions and additional probing comments, and to share their opinions with group members even if they disagreed. Focus groups were conducted by a white woman licensed clinical psychologist (S.B.) with experience working with dementia care-partners, and weekly supervision was provided by another white woman licensed clinical psychologist (A-M.V.). A white woman clinical research coordinator (J.B.) observed interviews to navigate technical difficulties, take field notes, and ensure engagement of focus group participants (e.g., asking probing questions, asking participants for additional input). Interviews were audio-recorded and transcribed verbatim (identifiers removed) using the TranscribeMe professional transcription service. Transcripts were not returned to participants for comment.

### Data Analytic Plan

We applied a hybrid deductive-inductive approach to thematic data analysis (Fereday & Muir-Cochrane, 2006). Specifically, we used *deductive* techniques in the sense that we drew from prior research on adjustment to ADRDs to develop our interview guide and initial codebook domains (McDermott et al., 2019; Oyebode & Parveen, 2019). The study team involved in the coding process (S.B., J.B., N.A.) began by developing familiarity with the data via interview transcripts and team members’ observations, and then identified preliminary codes (i.e., conceptual labels assigned to transcript excerpts such as “sources of emotional distress”). The coding team then *inductively* organized codes into overarching domains that were determined a-priori using deductive techniques. We coded the dataset using the Dedoose software package (version 9.0.17) (SocioCultural Research Consultants LLC, 2021) and systematically applied our final codebook across the dataset. All transcripts were coded independently by two members of the research team (J.B. and N.A.). Discrepancies were resolved through weekly hour-long discussions with the coders and senior author (S.B.), consistent with guidelines for team-based analysis (Giesen & Roeser, 2020).

We adopted a collaborative and iterative approach to data interpretation. Two members of the analysis team (A.H. and S.B.) organized findings within supraordinate themes, defined subthemes, and identified illustrative quotes in a step-wise process. First, we clustered findings within broader categories and identified initial subthemes and definitions. Then, we iteratively revised all extracted findings to improve clarity and reduce redundancy and overlap. We discussed our initial findings with the larger study team to ensure that the identified findings reflected the coded data and were clear to readers. Finally, our analysis team (A.H. and S.B.) selected representative quotes to describe identified themes. Participants did not provide feedback on the findings.

## RESULTS

### Sample Characteristics

Participant characteristics are presented in Table 1. Participants were recruited from Massachusetts General Hospital (*n*=12; 63.2%) and Mount Sinai Hospital (*n*=7; 36.8%) sites. Participants were predominantly female (*n*=14; 73.7%), white non-Hispanic or Latinx (*n*=15; 78.9%), and 41.5 years old (SD=10.1) on average. They had a range of professional backgrounds, including geriatrician (*n*=3; 17.6%), social worker (*n*=3; 17.6%), nurse/nurse practitioner (*n*=2; 11.8%); neurologist (*n*=3; 17.6%), geriatric psychiatrist (*n*=3; 17.6%), caregiver support director/coordinator (*n*=2; 11.8%), and neuropsychologist (*n*=1; 6%). Although the two caregiver support specialists did not have formal clinical training, they had undergone informal training and had practiced for more than 5 years providing direct one-on-one or group support to dementia caregivers and in some cases patients.

**Table 1:**
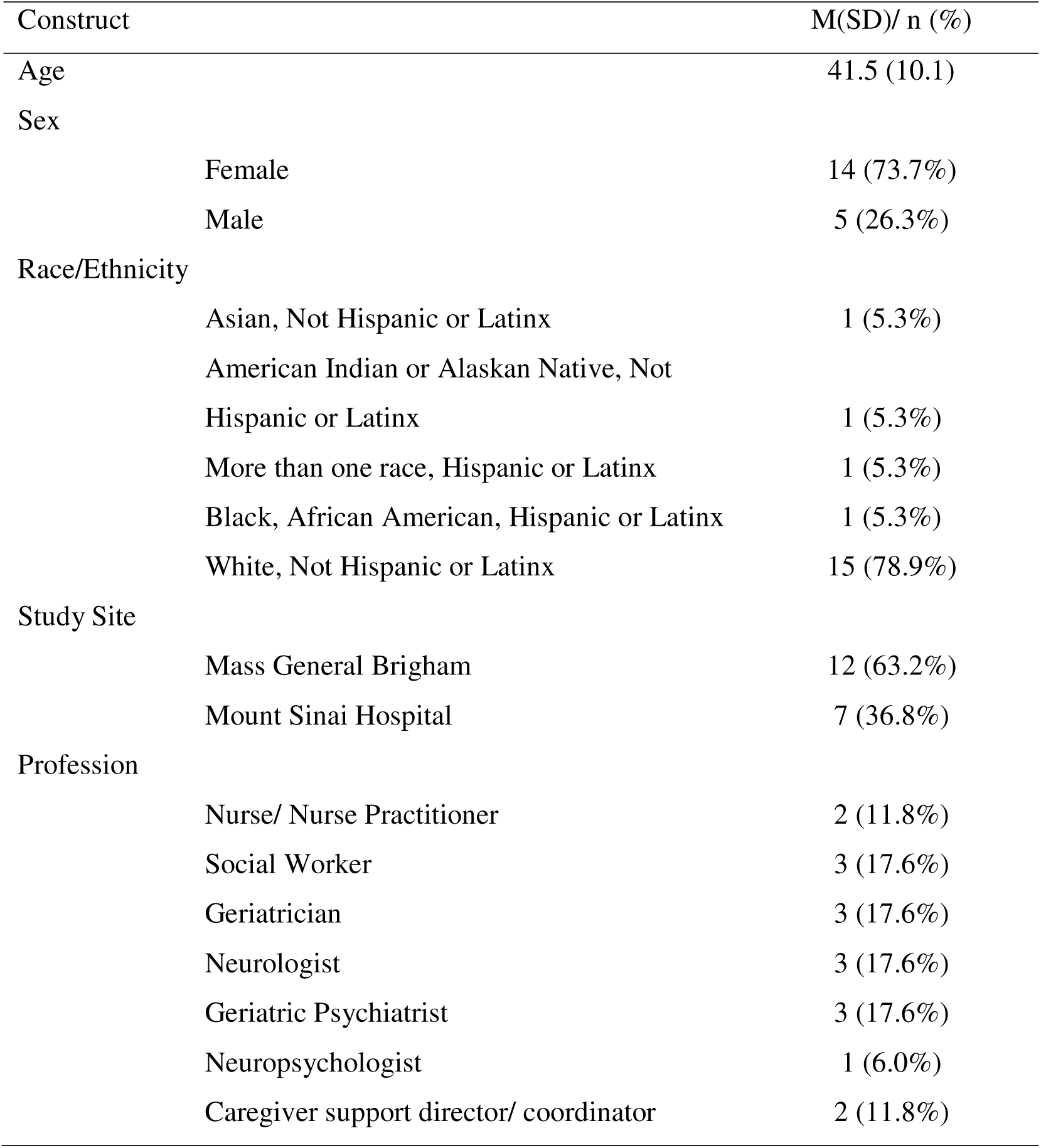
Participant Characteristics (n=19)

### Themes and Subthemes

We organized data in three deductive themes determined a priori. We inductively identified several sub-themes within the three overarching themes. Detailed descriptions of each theme are provided below. Table 2 provides illustrative quotations within each theme. We include the dementia care specialists’ hospital site and role with descriptive quotations to place quotations in relevant contexts.

**Table 2:**
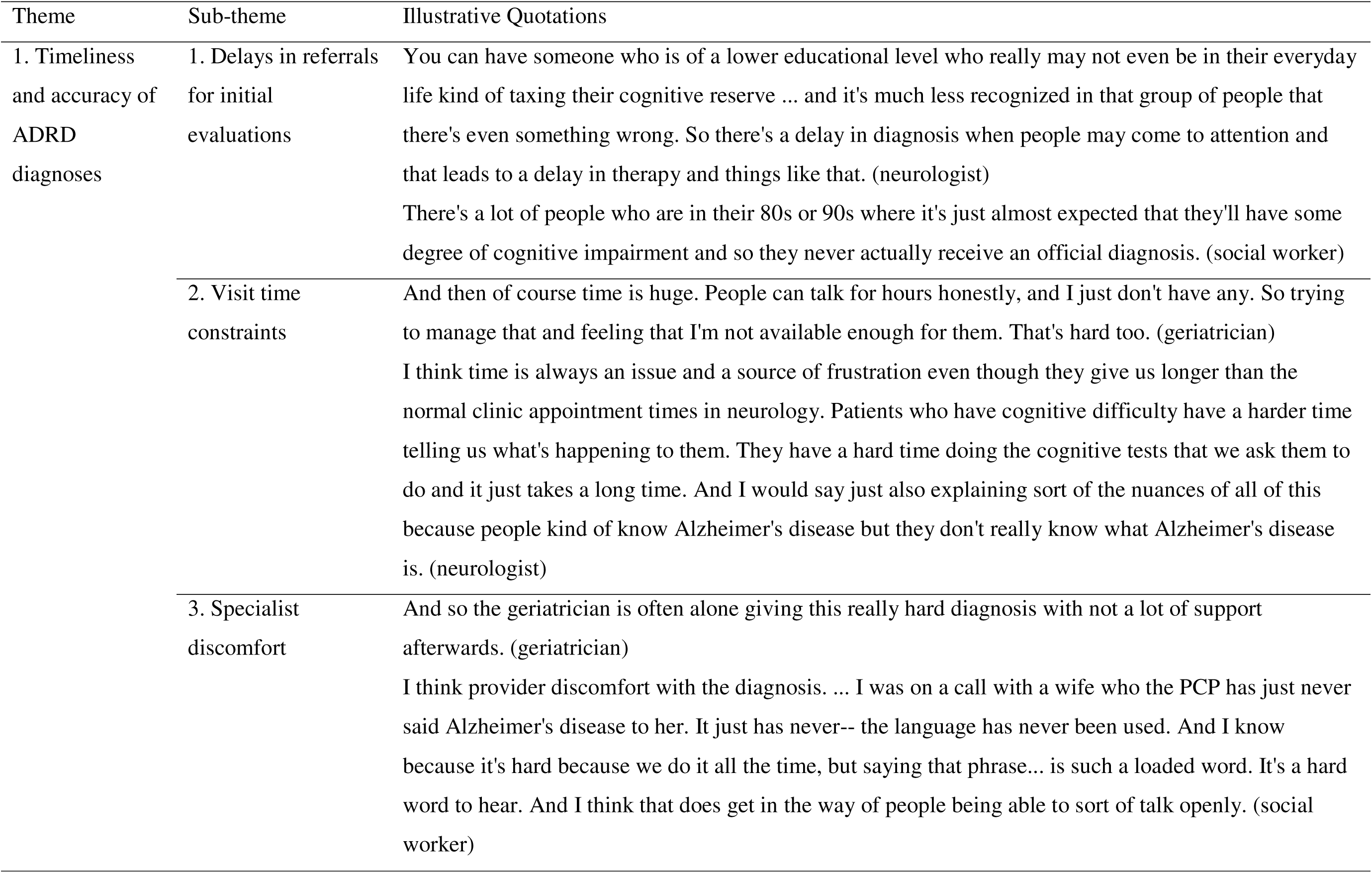
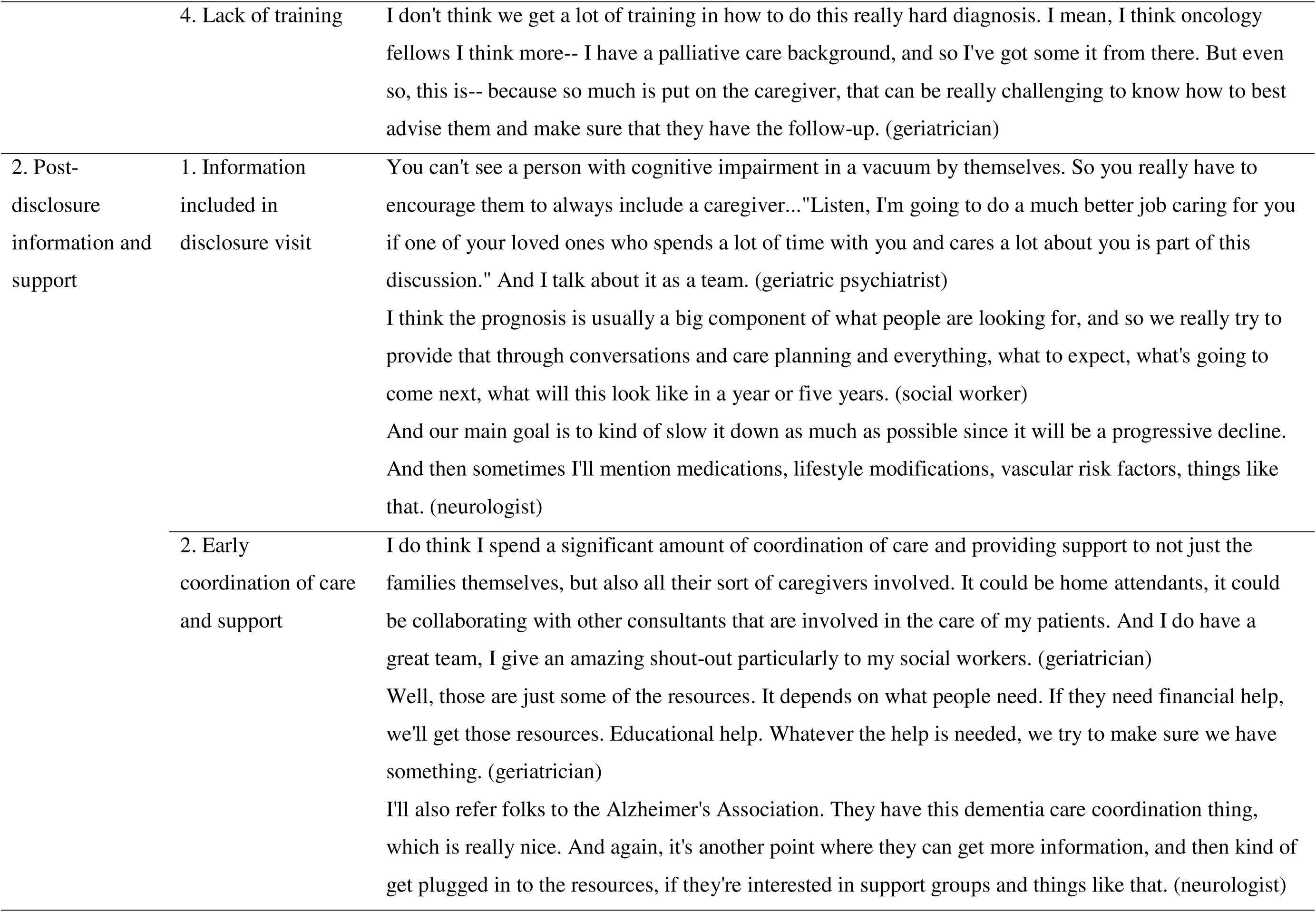
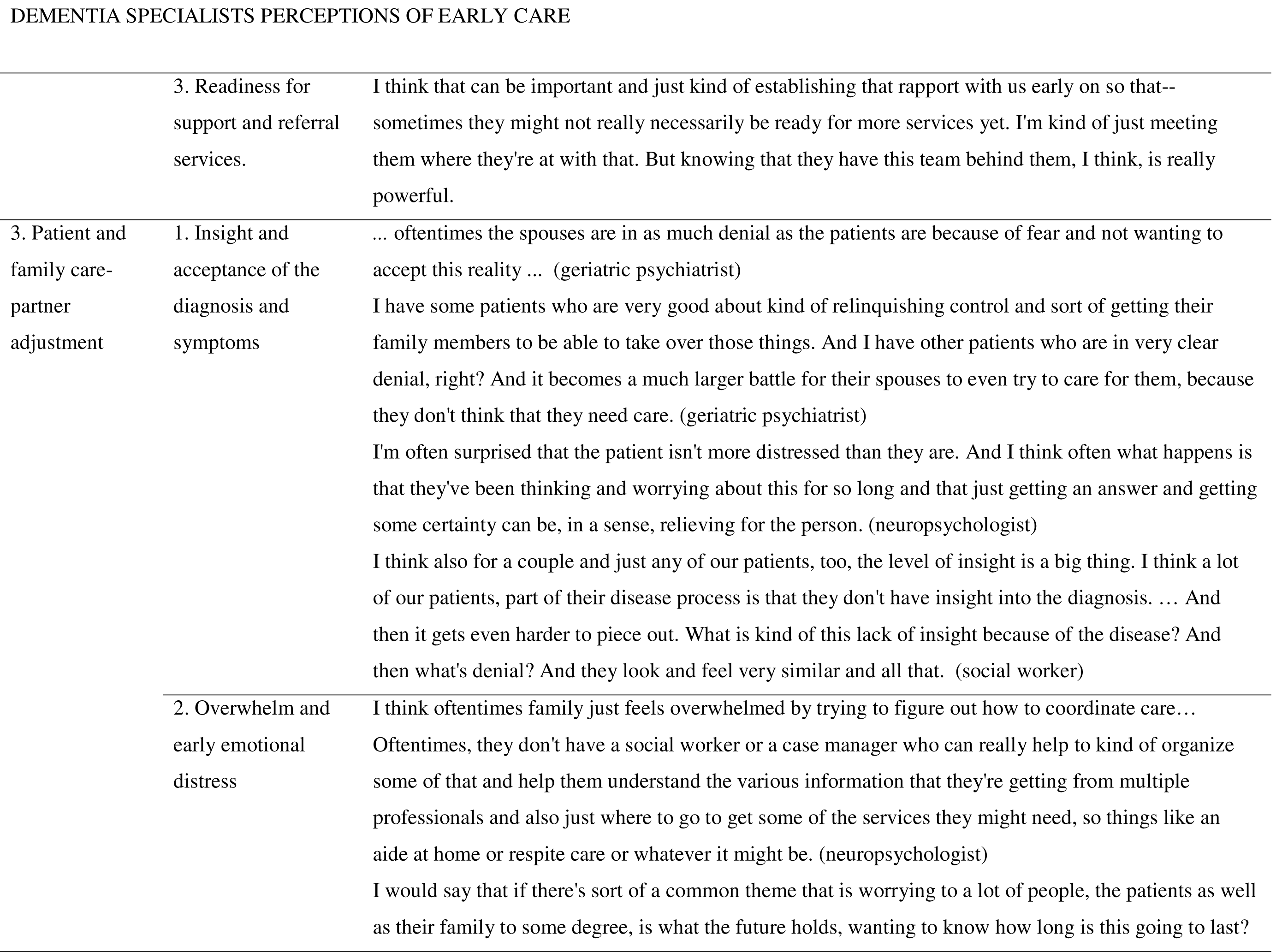
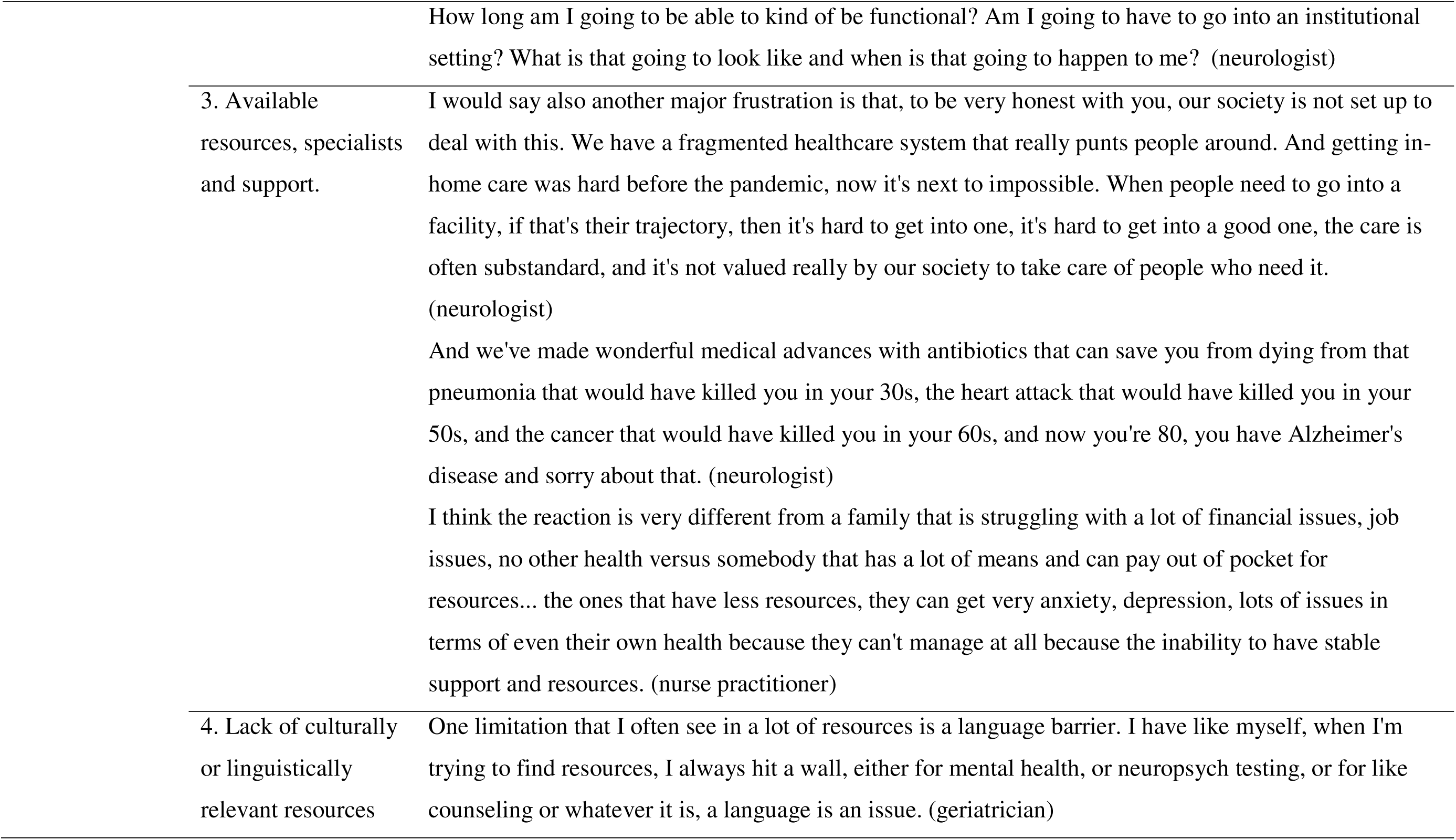
Description of Themes, Subthemes, and Illustrative Quotations by Study Domain.

### Theme 1: Timeliness and accuracy of ADRD diagnoses

We identified four sub-themes pertaining to dementia care specialists’ perspectives on timely and accurate diagnosis: (1) delays in referrals for initial evaluations; (2) visit time constraints; (3) dementia care specialist discomfort; and (4) lack of training.

Dementia care specialists felt that delays in referrals were related to patient, family, and specialist expectations about what is considered “normal aging,” and to patients’ education and employment background that could lead to symptoms being missed or misattributed to other causes (subtheme 1; delays in referrals for initial evaluations).

Specialists involved in diagnostic determinations also expressed frustration that their time with patients was significantly constrained by long waitlists and short patient visits, which impacted evaluations, the confidence of diagnoses, and the timing of disclosure visits (subtheme 2; visit time constraints). Specialists communicated that diagnostic accuracy often had to be balanced against clinical efficiency. Due to long waitlists, lack of service providers, and insufficient treatments, specialists described a hesitation between making a diagnosis early—and thereby facilitating greater time for adjustment and provision of resources—and the desire for more diagnostic certainty that is provided by multiple assessments and patient interactions to gather more complete information about a patient’s functioning (over time and across modalities).

Dementia care specialists noted that delays in making (i.e., formulating) and communicating (i.e., verbal/ written disclosure) diagnoses were also due to a lack of knowledge, training, and comfort in diagnosing ADRDs (subtheme 3; dementia care specialist discomfort and subtheme 4; lack of training). This was perceived to be the case for generalists but was also expressed by specialists in the study. Many specialists, including neurologists and geriatricians, emphasized their own or other colleagues’ discomfort in making an ADRD diagnosis. However, discomfort with diagnostic determination appeared to differ by participant profession and specialty. This was attributed to multiple factors including differences in levels of expertise and the presence or absence of sufficient information from referring clinicians to make a diagnosis without additional assessment. Specialists pointed specifically to the challenges faced by primary care providers, who are well-positioned to detect new symptoms, but may be even more uncomfortable making diagnostic determinations. All specialists interviewed were acutely aware of the gravity of providing this terminal diagnosis, and many expressed emotional discomfort in their role in communicating diagnostic information.

### Theme 2: Post-disclosure information and support

We identified three sub-themes pertaining to dementia care specialists’ experience sharing information about ADRD diagnoses and post-diagnosis support to dyads: (1) information included in disclosure visits; (2) early coordination of care and support; and (3) readiness for support and referral services.

Regarding disclosure visits (subtheme 1; information included in disclosure visits), specialists shared the importance of discussing collaborative care management with both patients and care-partners and working to give individuals and families prognostic information for near-and long-term care plans. The process of helping people “know what to expect,” was viewed as difficult for specialists given the challenges confirming a diagnosis and in recognition of the heterogeneous symptom trajectories across individuals. Many specialists also included a discussion of modifiable lifestyle factors within disclosure visits, to help patients maintain physical and cognitive health and potentially slow progression of symptoms where possible.

Some specialists also mentioned going to lengths to connect individuals and families to social, financial, legal, and community resources and caregiving assistance (subtheme 2; early coordination of care and support). Specialists emphasized the difficulty addressing the breadth of symptoms and care needs common to individuals with ADRDs and to coach care partners to provide support where appropriate. Many discussed their efforts to tailor services to patients’ and families’ needs and to coordinate care among all caregivers involved.

Dementia care specialists discussed their process of finding the balance of early support provision and respecting patients’ and families’ readiness for services (subtheme 3; readiness for support and referral services). Many specialists reflected that families were not ready to receive services in the immediate post-diagnosis period as they were experiencing feelings of shock, grief, stress, and may have been experiencing denial. Social workers were consistently identified by other specialists as critical in the early period, however, were commonly described as difficult to access. Many used organizations like the Alzheimer’s Association to fill critical gaps in providing psychosocial resources.

### Theme 3: Patient and family care-partner adjustment

We identified four subthemes pertaining to dementia care specialists’ impressions of factors impacting patient and care-partner adjustment: (1) insight and acceptance of the diagnosis and symptoms; (2) overwhelm and early emotional distress; (3) available resources, specialists, and support; and (4) lack of culturally or linguistically relevant resources.

Specialists mentioned the importance of assessing and promoting understanding and appreciation of the diagnosis and symptoms in both patients and care-partners at the time of diagnosis and at follow-up visits (subtheme 1; insight and acceptance of the diagnosis and symptoms). Many acknowledged the tendency for patients and family care-partners to deny their reality in the period shortly after diagnosis amid overwhelming emotions. They felt that denial was an important barrier to open and collaborative communication and positive adjustment within families. A lack of insight into symptoms was also identified as a key barrier and was reportedly more common for those diagnosed later in the clinical course. Specialists expressed that distinguishing between denial and lack of insight could be difficult yet was extremely important for determining their approach to care.

There was consensus that experiences of feeling overwhelmed and early emotional distress were common for individuals and families and sometimes persisted long after the diagnosis (subtheme 2; overwhelm and early emotional distress). Families expressed significant fear and worry about the future and how the progressive decline would evolve. Care-partners were often not surprised by the diagnosis but expressed a fear of navigating long-term care needs (e.g., allied health care, in-home support, respite services, and navigating insurance and benefits) once it was confirmed. Role changes in the family unit led to changes in identity and disruption of familiar dynamics, which was a common source of multiple negative emotions especially for those in denial of their diagnosis (e.g., grief, frustration, worry, hopelessness). Specialists reflected on the importance of fluidity and flexibility in navigating role change and preserving some aspects of previously valued roles or independence with support and structure where possible. Specialists noted that feelings of being overwhelmed in either patients or family care-partners was a barrier to positive adjustment—as overwhelmed individuals often coped with avoidance or denial, which served as barriers to collaborative coping.

In addition to these challenges, specialists emphasized the lack of available resources, specialists, and ongoing supports (subtheme 3; available resources, specialists, and support) as factors impacting distress and adjustment for most families. Specialists also acknowledged the limitations of existing services and resources for a diverse range of patients in terms of language and culture (subtheme 4; lack of culturally or linguistically relevant resources). They noted that the field’s understanding of early ADRD symptoms (e.g., cognition, changes in cognition and what constitutes a problematic change in cognition) is fundamentally linked to language and culture, which impacts the timeliness and accuracy of diagnosis and continuum of care. For example, there were language and cultural barriers to receiving a neuropsychological assessment, which is a key component of comprehensive ADRD diagnostic work-up. They also commonly acknowledged difficulties feeling prepared to work with patients and families with different backgrounds and identities (e.g., LGBTQ+ families), and expressed a desire for more training to promote culturally relevant care.

## DISCUSSION AND IMPLICATIONS

The present study had the primary objective of characterizing dementia care specialist perspectives of ADRD diagnosis, post-diagnosis support, and early dyadic adjustment. We used a hybrid deductive-inductive approach to thematic analysis of focus groups and individual qualitative interviews with a diverse pool of dementia care specialists (geriatricians, social workers, nurses/nurse practitioners, neurologists, geriatric psychiatrists, caregiver support coordinators, and neuropsychologists) from two large academic medical centers. We leveraged theory and prior research to inductively identify subthemes within three a-priori determined themes that explored factors impacting accurate and timely diagnosis, diagnostic disclosure and early support provision, and factors impacting dyadic adjustment. These themes are closely inter-related, with timely and accurate diagnosis facilitating earlier and more effective post-diagnosis support and consequently promoting adjustment.

Dementia care specialists in our study agreed that barriers to timely diagnosis were frequent and difficult to navigate (Parker et al., 2020). Our study echoes previous survey data showing that specialists find it difficult to differentiate ADRDs from normal cognitive aging (Bradford et al., 2009) and suggests greater training is required to confidently differentiate normal aging from early symptoms of dementia in the early stage of illness progression (Bernstein et al., 2019). Specialists also identified long waitlists and visit time constraints for typical appointments as challenges that impacted the diagnostic evaluation process and the time they could allocate to diagnostic disclosure in patient visits. This has been raised in several other studies over many years (Gauthier et al., 2022), though some improvements have been made with recent innovations (e.g., the UCLA ADC program) that have not been disseminated to other large academic medical centers (Reuben et al., 2022). Long wait lists, visit time constraints, and other barriers to patient-centered disclosure practices are important to consider, as reaction to the diagnosis and its ramifications (e.g. recommendations to cease driving, relocate to a supportive living environment) can be so overwhelming that it interferes with the cognitive intake of the information disclosed (Aminzadeh et al., 2007; Bailey et al., 2019). Given their constraints and challenges, it is not surprising that specialists reflected introspectively on their own discomfort in providing a diagnosis. This is particularly notable given that our sample comprised specialists and coordinators with expert knowledge and experience in dementia clinical care at well-resourced academic medical centers.

We also identified novel themes surrounding specialists’ impressions of the practical logistics of diagnostic disclosure and post-diagnosis support. Specialists cited preferences that disclosure happen with care partners present whenever possible (Gauthier et al., 2022). During disclosure visits, specialists felt that highlighting lifestyle factors that could slow cognitive decline was important to promote positive health management and inspire hope. Specialists’ also felt it was important to be open and realistic about patients’ prognosis. These preferences are all consistent with recent suggestions for optimizing diagnosis disclosure and post-diagnosis care (Armstrong et al., 2024; Gauthier et al., 2022), which indicates that specialists at large academic medical centers are practicing in manners consistent with recommendations despite some enduring structural barrriers.

Finally, dementia care specialists highlighted the critical role of emotional distress as a key barrier to early adjustment for patients and care-partners. The causes for this were multi-factorial, but specialists emphasized the burden of care coordination, fears for the future, and changes in family roles and identity. This aligns with patients’ perspectives from previous research that have also described the distress and grief associated with loss of valued life roles (both professional and familial) and changes to their stable sense of identity (Aminzadeh et al., 2007; Petty et al., 2018). They noted that patients’ and family members’ levels of insight and acceptance of the diagnosis impacted early distress, as well as the availability of specialists and culturally or linguistically appropriate care. Our findings highlight specialists’ awareness of the many and diverse factors linked to early adjustment following diagnostic disclosure.

### Clinical Implications

Our findings are particularly timely considering the recent advances in health-systems improvements in dementia care coordination and recognition of the difficulties in implementing such innovations. Taken together, our findings highlight continued gaps in care and ways of improving services for patients and care partners early after ADRD diagnosis. Specifically, specialists lack of training in diagnostic disclosure has been highlighted in many reports over many years (Bailey et al., 2019; Bradford et al., 2009; Dubois et al., 2016; Yates et al., 2021), suggesting there has been little progress on this issue. Additional training should include how cognitive impairments may impact patient comprehension of diagnosis, and how to manage specialists own emotional distress (Bailey et al., 2019; Yates et al., 2021). Specialists may benefit from training in semi-structured approaches to diagnostic disclosure such as SPIKES (Peixoto et al., 2020) or the serious illness communication approach (Goyal et al., 2023). More education is also needed to improve the cultural and linguistic appropriateness of diagnostic evaluations, disclosure, and post-diagnosis care (*2023 Alzheimer’s Disease Facts and Figures*, 2023; Lin et al., 2021; Yates et al., 2021).

Visit structures in specialty and generalist clinics should facilitate diagnostic disclosure occurring over multiple longer visits (Aminzadeh et al., 2007; L. Robinson et al., 2005; Yates et al., 2021). This would allow for more opportunities to impart the same information, or present smaller amounts of information more than once, which could help patients and families to remember the diagnosis and its implications and provide space to explore the emotional impact of the diagnosis (Yates et al., 2021). Adequate information provision is critical in this early period as developing a greater understanding of what is happening helps to regain sense of control and facilitates coping (L. Robinson et al., 2005). While these potential benefits are important, increases in the workforce are necessary to avoid increased wait times for visits, as well as broader systems level changes such as revised reimbursement structures to allow billing for complex care hours. Given that primary care providers commonly express that specialists (e.g., behavioral neurologists, geriatricians, neuropsychologists) are better to assess and diagnose patients (Yates et al., 2021), healthcare systems should consider adopting strategies that provide generalists with support and guidance to develop their knowledge and confidence in dementia care.

The need for early support and the critical role of emotional distress in adjustment to ADRDs emphasized by specialists in our study highlights the necessity of psychosocial interventions that begin early in the diagnostic journey. The post-diagnostic period provides a critical window for interventions when levels of burden are low for families, and persons living with dementia are more able to meaningfully participate in their care (de Vugt & Verhey, 2013). Psychosocial interventions should focus on adjustment, adaptive and positive coping, and psychoeducation to facilitate acceptance of future transitions and care roles (Aminzadeh et al., 2007; de Vugt & Verhey, 2013). Given the demands on dementia care specialists and lack of available resources at present, approaches such as dyadic interventions that simultaneously involve patients and care-partners may be a more efficient and effective way of improving post-diagnosis care (Bannon, Brewer, Ahmad, et al., 2023; Vranceanu et al., 2022). Evidence suggests that patients and care-partners are interested in such interventions (Byszewski et al., 2007; Parker et al., 2020), and dyadic interventions for later stages of ADRDs show positive effects on relationships and have a greater impact on long-term adjustment to ADRDs than individually-focused approaches (Gellert et al., 2018; Moon & Adams, 2013). Despite this need, there are currently no established psychosocial interventions available at the time of diagnosis for people living with ADRDs or their care partners (Parker et al., 2020).

### Study Limitations

We identified several potential limitations. We recruited a multidisciplinary group of medical dementia care specialists from two academic medical center hospital systems in the Boston and New York metropolitan areas, and it is possible that specialists in care settings with a different clinical infrastructure or geographic regions would have different perspectives. In addition, we used convenience sampling through established clinics and department listservs, which may have led to our sample being a more engaged group relative to those who did not respond to study invitations. Finally, our sample largely comprised of White women, and it is likely that specialists from different backgrounds would offer additional perspectives on existing services and potential avenues for refinement.

### Conclusions

Our study sought to characterize dementia care specialists’ impressions of factors impacting ADRD diagnoses and post-diagnosis support. Specialist highlighted the various challenges with: (1) timely and accurate diagnosis, (2) diagnostic disclosure and the provision of post-diagnosis support, and (3) dyadic adjustment after diagnosis. Results from this study have the potential to inform plans for post-diagnosis clinical care for individuals, couples, and families, with implications for other neurodegenerative diseases.

## Supporting information

Table 1: Participant Characteristics

Table 2: Description of Themes, Subthemes, and Illustrative Quotations by Study Domain

Supplemental Material: Interview Script

Supplemental Material: Final Codebook

## Acknowledgements

We would like to thank the members of the MGH CHOIR lab members and the MGH “K club” led by Dr. Ana-Maria Vranceanu for their feedback on proposed focus group procedures.

## Data Availability Statement

Data collection and analysis instruments are available in the Supplemental Materials. The data sets generated during and/or analyzed during the current study are available to qualified researchers from the corresponding author on reasonable request.

## Supplemental Material: Interview Script

### Warm Up

To start, could each of you please tell me your name, the hospital you work in, your professional position, and the type of care you provide for persons living with ADRDs?

[*Note: Ensure that each participate provides responses.*]

### Theme: Clinical Care Early after ADRD Diagnosis

We’d like to learn more about the typical clinical care persons living with ADRD receive before and after ADRD diagnoses.

How would you describe the experience of delivering an ADRD diagnosis to patients and their caregivers?

Probe: Typical clinic flow to receive a diagnosis
Probe: Who attends appointments
**Probe: How information is delivered**
**Probe: Typical follow-up visits and support** Probe: Points of contact
Probe: Providers’ challenges delivering diagnosis or communicating with couples
Probe: Providing referrals to support groups, individual counseling, couples therapy

What do you notice regarding emotional distress (depression, anxiety, stress) in couples after diagnoses?

Probe: How common within the period 1-2 months after diagnoses
Probe: Observations on any contextual factors that impact distress
Probe: **Mental health resources available to patients**
**Probe:** Ideal scenario for addressing distress and mental health concerns after diagnoses

**What resources are available for common challenges that patients and caregivers experience <u>immediately after diagnosis?</u>**

Probe: Emotional support
Probe: Understanding the diagnosis and next steps
Probe: Resources for help managing symptoms and adjusting to care-partnership
Probe: Communicating about difficult emotions and relationship changes
Probe: Planning for the future

**What, if any, are the sources of stress for you (as the provider) working in this setting?**

## Supplemental Material: Final Codebook

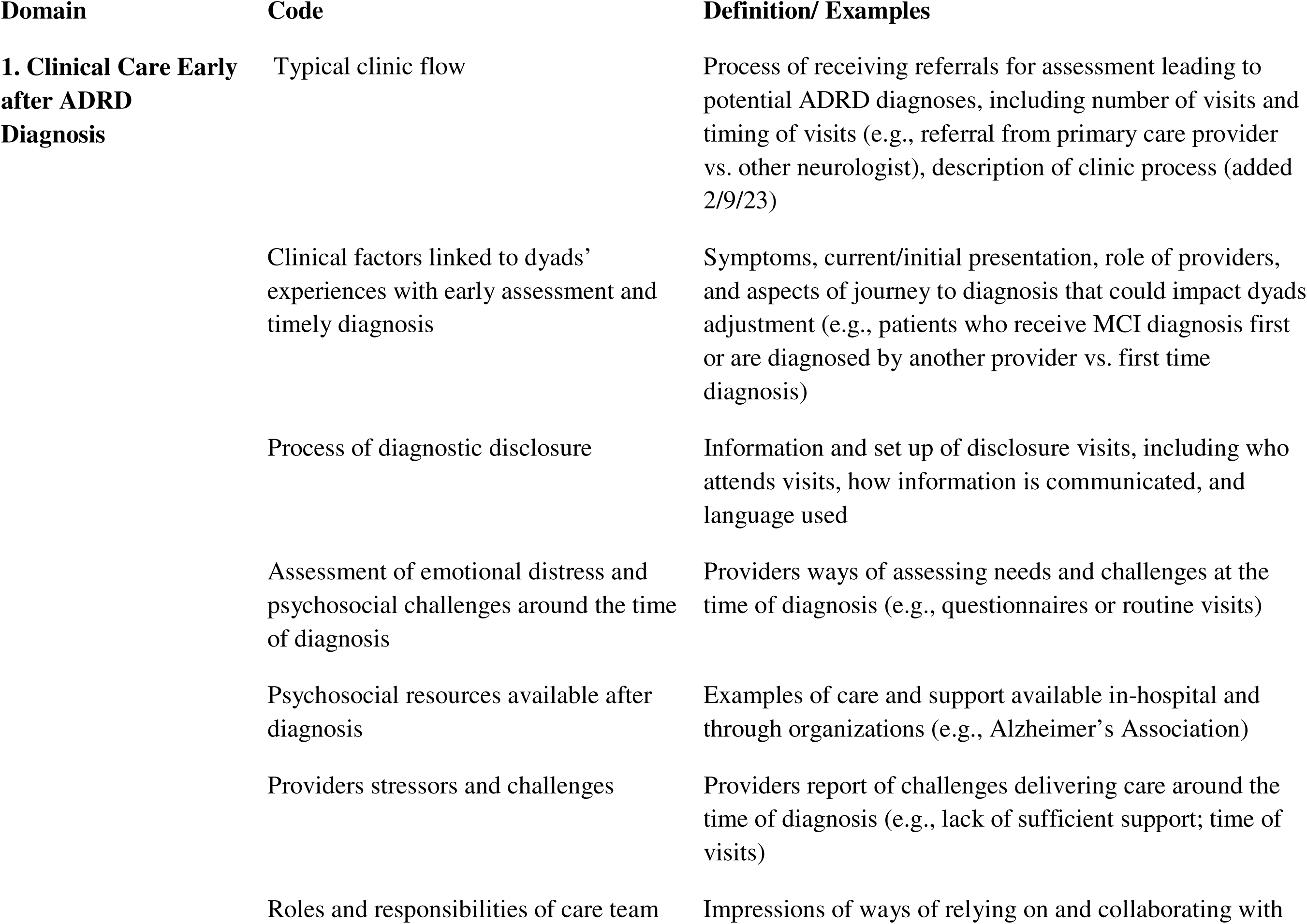
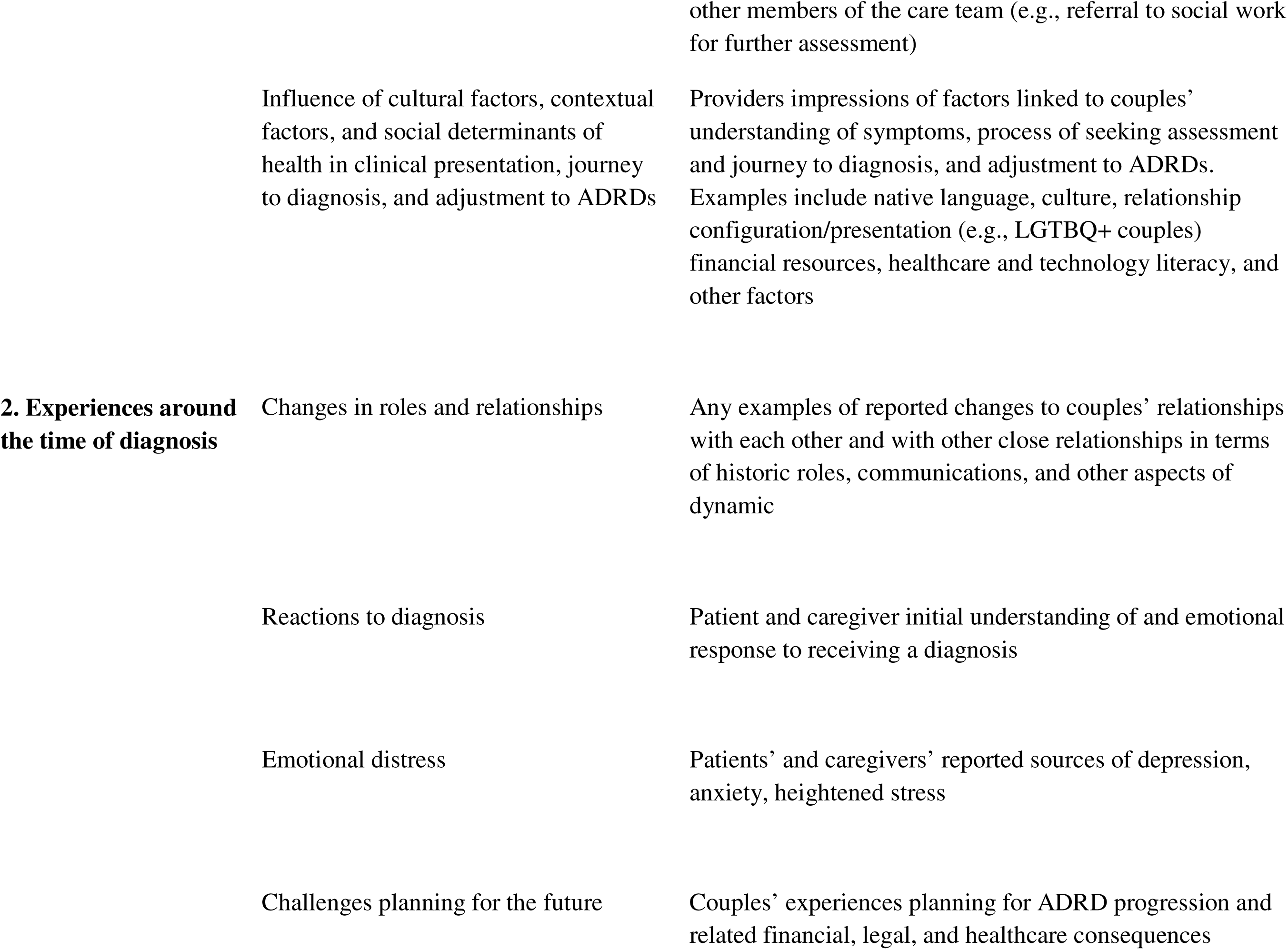
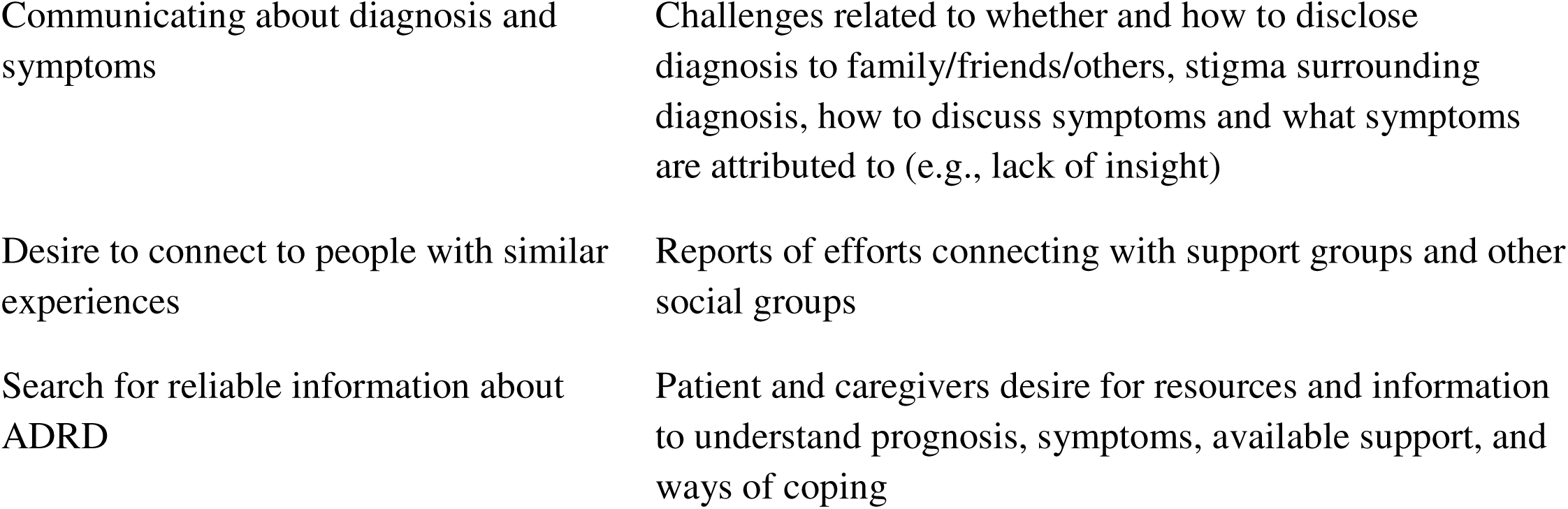

