## Supplementary material for "Dementia Care Specialists Perspectives of Diagnosis and Early Psychosocial Care: A Qualitative Analysis of Focus Groups in Two Large Academic Medical Centers": Table 1: Participant Characteristics

**Table 1: Participant Characteristics (n=19)**

**Table 1**

*Participant Characteristics (n=19)*

| Construct |  |  | M(SD)/ n (%) |
| --- | --- | --- | --- |
| Age |  |  | 41.5 (10.1) |
| Sex |  |  |  |
|  | Female |  | 14 (73.7%) |
|  | Male |  | 5 (26.3%) |
| Race/Ethnicity | |  |  |
|  | Asian, Not Hispanic or Latinx | | 1 (5.3%) |
|  | American Indian or Alaskan Native, Not Hispanic or Latinx | | 1 (5.3%) |
|  | More than one race, Hispanic or Latinx | | 1 (5.3%) |
|  | Black, African American, Hispanic or Latinx | | 1 (5.3%) |
|  | White, Not Hispanic or Latinx | | 15 (78.9%) |
| Study Site | | |  |
|  | Mass General Brigham | | 12 (63.2%) |
|  | Mount Sinai Hospital |  | 7 (36.8%) |
| Profession | | |  |
|  | Nurse/ Nurse Practitioner | | 2 (11.8%) |
|  | Social Worker |  | 3 (17.6%) |
|  | Geriatrician |  | 3 (17.6%) |
|  | Neurologist |  | 3 (17.6%) |
|  | Geriatric Psychiatrist |  | 3 (17.6%) |
|  | Neuropsychologist |  | 1 (6.0%) |
|  | Caregiver support director/ coordinator |  | 2 (11.8%) |
