## Supplementary material for "Dementia Care Specialists Perspectives of Diagnosis and Early Psychosocial Care: A Qualitative Analysis of Focus Groups in Two Large Academic Medical Centers": Table 2: Description of Themes, Subthemes, and Illustrative Quotations by Study Domain

**Table 2**

*Description of Themes, Subthemes, and Illustrative Quotations by Study Domain*

| Theme | Sub-theme | Illustrative Quotations |
| --- | --- | --- |
| 1. Timeliness and accuracy of ADRD diagnoses | 1. Delays in referrals for initial evaluations | You can have someone who is of a lower educational level who really may not even be in their everyday life kind of taxing their cognitive reserve ... and it's much less recognized in that group of people that there's even something wrong. So there's a delay in diagnosis when people may come to attention and that leads to a delay in therapy and things like that. (neurologist)  There's a lot of people who are in their 80s or 90s where it's just almost expected that they'll have some degree of cognitive impairment and so they never actually receive an official diagnosis. (social worker) |
|  | 2. Visit time constraints | And then of course time is huge. People can talk for hours honestly, and I just don't have any. So trying to manage that and feeling that I'm not available enough for them. That's hard too. (geriatrician)  I think time is always an issue and a source of frustration even though they give us longer than the normal clinic appointment times in neurology. Patients who have cognitive difficulty have a harder time telling us what's happening to them. They have a hard time doing the cognitive tests that we ask them to do and it just takes a long time. And I would say just also explaining sort of the nuances of all of this because people kind of know Alzheimer's disease but they don't really know what Alzheimer's disease is. (neurologist) |
|  | 3. Specialist discomfort | And so the geriatrician is often alone giving this really hard diagnosis with not a lot of support afterwards. (geriatrician)  I think provider discomfort with the diagnosis. ... I was on a call with a wife who the PCP has just never said Alzheimer's disease to her. It just has never-- the language has never been used. And I know because it's hard because we do it all the time, but saying that phrase... is such a loaded word. It's a hard word to hear. And I think that does get in the way of people being able to sort of talk openly. (social worker) |
|  | 4. Lack of training | I don't think we get a lot of training in how to do this really hard diagnosis. I mean, I think oncology fellows I think more-- I have a palliative care background, and so I've got some it from there. But even so, this is-- because so much is put on the caregiver, that can be really challenging to know how to best advise them and make sure that they have the follow-up. (geriatrician) |
| 2. Post-disclosure information and support | 1. Information included in disclosure visit | You can't see a person with cognitive impairment in a vacuum by themselves. So you really have to encourage them to always include a caregiver..."Listen, I'm going to do a much better job caring for you if one of your loved ones who spends a lot of time with you and cares a lot about you is part of this discussion." And I talk about it as a team. (geriatric psychiatrist)  I think the prognosis is usually a big component of what people are looking for, and so we really try to provide that through conversations and care planning and everything, what to expect, what's going to come next, what will this look like in a year or five years. (social worker)  And our main goal is to kind of slow it down as much as possible since it will be a progressive decline. And then sometimes I'll mention medications, lifestyle modifications, vascular risk factors, things like that. (neurologist) |
|  | 2. Early coordination of care and support | I do think I spend a significant amount of coordination of care and providing support to not just the families themselves, but also all their sort of caregivers involved. It could be home attendants, it could be collaborating with other consultants that are involved in the care of my patients. And I do have a great team, I give an amazing shout-out particularly to my social workers. (geriatrician)  Well, those are just some of the resources. It depends on what people need. If they need financial help, we'll get those resources. Educational help. Whatever the help is needed, we try to make sure we have something. (geriatrician)  I'll also refer folks to the Alzheimer's Association. They have this dementia care coordination thing, which is really nice. And again, it's another point where they can get more information, and then kind of get plugged in to the resources, if they're interested in support groups and things like that. (neurologist) |
|  | 3. Readiness for support and referral services. | I think that can be important and just kind of establishing that rapport with us early on so that-- sometimes they might not really necessarily be ready for more services yet. I'm kind of just meeting them where they're at with that. But knowing that they have this team behind them, I think, is really powerful. |
| 3. Patient and family care-partner adjustment | 1. Insight and acceptance of the diagnosis and symptoms | *...* oftentimes the spouses are in as much denial as the patients are because of fear and not wanting to accept this reality ... (geriatric psychiatrist)  I have some patients who are very good about kind of relinquishing control and sort of getting their family members to be able to take over those things. And I have other patients who are in very clear denial, right? And it becomes a much larger battle for their spouses to even try to care for them, because they don't think that they need care. (geriatric psychiatrist)  I'm often surprised that the patient isn't more distressed than they are. And I think often what happens is that they've been thinking and worrying about this for so long and that just getting an answer and getting some certainty can be, in a sense, relieving for the person. (neuropsychologist)  I think also for a couple and just any of our patients, too, the level of insight is a big thing. I think a lot of our patients, part of their disease process is that they don't have insight into the diagnosis. … And then it gets even harder to piece out. What is kind of this lack of insight because of the disease? And then what's denial? And they look and feel very similar and all that. (social worker) |
|  | 2. Overwhelm and early emotional distress | I think oftentimes family just feels overwhelmed by trying to figure out how to coordinate care… Oftentimes, they don't have a social worker or a case manager who can really help to kind of organize some of that and help them understand the various information that they're getting from multiple professionals and also just where to go to get some of the services they might need, so things like an aide at home or respite care or whatever it might be. (neuropsychologist)  I would say that if there's sort of a common theme that is worrying to a lot of people, the patients as well as their family to some degree, is what the future holds, wanting to know how long is this going to last? How long am I going to be able to kind of be functional? Am I going to have to go into an institutional setting? What is that going to look like and when is that going to happen to me? (neurologist) |
|  | 3. Available resources, specialists and support. | I would say also another major frustration is that, to be very honest with you, our society is not set up to deal with this. We have a fragmented healthcare system that really punts people around. And getting in-home care was hard before the pandemic, now it's next to impossible. When people need to go into a facility, if that's their trajectory, then it's hard to get into one, it's hard to get into a good one, the care is often substandard, and it's not valued really by our society to take care of people who need it. (neurologist)  And we've made wonderful medical advances with antibiotics that can save you from dying from that pneumonia that would have killed you in your 30s, the heart attack that would have killed you in your 50s, and the cancer that would have killed you in your 60s, and now you're 80, you have Alzheimer's disease and sorry about that. (neurologist)  I think the reaction is very different from a family that is struggling with a lot of financial issues, job issues, no other health versus somebody that has a lot of means and can pay out of pocket for resources... the ones that have less resources, they can get very anxiety, depression, lots of issues in terms of even their own health because they can't manage at all because the inability to have stable support and resources. (nurse practitioner) |
|  | 4. Lack of culturally or linguistically relevant resources | One limitation that I often see in a lot of resources is a language barrier. I have like myself, when I'm trying to find resources, I always hit a wall, either for mental health, or neuropsych testing, or for like counseling or whatever it is, a language is an issue. (geriatrician) |
