## Supplemental Material: Final Codebook for "Dementia Care Specialists Perspectives of Diagnosis and Early Psychosocial Care: A Qualitative Analysis of Focus Groups in Two Large Academic Medical Centers"

| **Domain** | **Code** | **Definition/ Examples** |
| --- | --- | --- |
| **1. Clinical Care Early after ADRD Diagnosis** | Typical clinic flow | Process of receiving referrals for assessment leading to potential ADRD diagnoses, including number of visits and timing of visits (e.g., referral from primary care provider vs. other neurologist), description of clinic process (added 2/9/23) |
|  | Clinical factors linked to dyads’ experiences with early assessment and timely diagnosis | Symptoms, current/initial presentation, role of providers, and aspects of journey to diagnosis that could impact dyads adjustment (e.g., patients who receive MCI diagnosis first or are diagnosed by another provider vs. first time diagnosis) |
|  | Process of diagnostic disclosure | Information and set up of disclosure visits, including who attends visits, how information is communicated, and language used |
|  | Assessment of emotional distress and psychosocial challenges around the time of diagnosis | Providers ways of assessing needs and challenges at the time of diagnosis (e.g., questionnaires or routine visits) |
|  | Psychosocial resources available after diagnosis | Examples of care and support available in-hospital and through organizations (e.g., Alzheimer’s Association) |
|  | Providers stressors and challenges | Providers report of challenges delivering care around the time of diagnosis (e.g., lack of sufficient support; time of visits) |
|  | Roles and responsibilities of care team | Impressions of ways of relying on and collaborating with other members of the care team (e.g., referral to social work for further assessment) |
|  | Influence of cultural factors, contextual factors, and social determinants of health in clinical presentation, journey to diagnosis, and adjustment to ADRDs | Providers impressions of factors linked to couples’ understanding of symptoms, process of seeking assessment and journey to diagnosis, and adjustment to ADRDs. Examples include native language, culture, relationship configuration/presentation (e.g., LGTBQ+ couples) financial resources, healthcare and technology literacy, and other factors |
| **2. Experiences around the time of diagnosis** | Changes in roles and relationships | Any examples of reported changes to couples’ relationships with each other and with other close relationships in terms of historic roles, communications, and other aspects of dynamic |
|  | Reactions to diagnosis | Patient and caregiver initial understanding of and emotional response to receiving a diagnosis |
|  | Emotional distress | Patients’ and caregivers’ reported sources of depression, anxiety, heightened stress |
|  | Challenges planning for the future | Couples’ experiences planning for ADRD progression and related financial, legal, and healthcare consequences |
|  | Communicating about diagnosis and symptoms | Challenges related to whether and how to disclose diagnosis to family/friends/others, stigma surrounding diagnosis, how to discuss symptoms and what symptoms are attributed to (e.g., lack of insight) |
|  | Desire to connect to people with similar experiences | Reports of efforts connecting with support groups and other social groups |
|  | Search for reliable information about ADRD | Patient and caregivers desire for resources and information to understand prognosis, symptoms, available support, and ways of coping |
